# Sexual Orientation Experiences of Discrimination: Dimensionality, internal consistency, and nomological validity in a diverse Colombian population

**DOI:** 10.1101/2022.02.06.22269563

**Authors:** Yeison David Gallo-Barrera, Adalberto Campo-Arias

## Abstract

The study aimed to determine the dimensionality, internal consistency, and nomological validity of the Sexual Orientation Experiences of Discrimination (SOEOD-9) among lesbian, gay, bisexual, and queer people in Santa Marta, Colombia. The study included 303 individuals between 18 and 70 years old. Participants completed the SOEOD-9. Dimensionality was explored using confirmatory (CFA) factor analysis; internal consistency with Cronbach’s alpha and McDonald’s omega; and nomological validity with Student’s test. The SOEOD-9 presented two factors that explained 51.6% of the variance and high internal consistency. Nevertheless, the CFA showed poor indicators of goodness-of-fit for the two-dimensional solution. A five-item version (SOEOD-5) improved overall performance (dimensionality, internal consistency, and nomological validity). In conclusion, the SOEOD-9 shows a two-factor internal structure with poor goodness-of-fit indicators and acceptable internal consistency and nomological validity. The SOEOD-5 presents better global indicators of dimensionality, internal consistency, and nomological validity.

---

Stigma, prejudice, stereotype, and discrimination are phenomena studied by psychology, anthropology, and sociology (Hendry, 2016; Stangor, 2016), mainly based on attribution theory (Hernández & López, 2011). In recent years, these terms were unified in a theoretical construct called the “stigma-discrimination complex,” a complex, indivisible, spiral sequence that begins with stigma, continues as prejudice, becomes a stereotype, and ends in discrimination (Campo-Arias & Herazo, 2014).

In this complex, social stigma is understood as a character with a negative connotation, which appears as a result of social interactions when the individual does not satisfy social expectations (Campo-Arias & Herazo, 2013; Goffman, 1963). Stigma gives way to prejudice when valuation is marked as undesirable; it is a value judgment with a solid emotional base that supports unfavorable attitudes towards a social group, which are manifested in negative attitudes towards members who belong to undesirable and devalued social categories (Allport, 1954; Campo-Arias & Herazo, 2015; Ramírez et al., 2016). A stereotype is a set of qualities, attributes, or specific traits attributed to the social group by which the group is homogenized and qualified (Campo-Arias & Herazo, 2014; Lippman, 1991). Stigma, prejudices, and stereotypes materialize in discrimination, which encompasses all behaviors, subtle or overt, of exclusion and systematic violence against groups that disturb norms, values, and social well-being according to the hegemonic group or that holds the social, political, or economic power (Awad & Rackley, 2017; López et al., 2008).

The stigma-discrimination complex can be presented against any social group. However, the most studied fields correspond to groups discriminated by race, ethnic origin, gender, religion, and sexual orientation (Carter et al., 2017; Ghumman & Ryan, 2018; Heilman & Caleo, 2018; Jones et al., 2017; Sugarman et al., 2018; Zschirnt & Ruedin, 2016). The experiences of discrimination, and more if they are repetitive, constitute a risk factor for the physical and mental health of the discriminated groups (Carter et al., 2019; De Freitas et al., 2018; Levefor et al., 2020; Semlyen et al., 2016). The stigma-discrimination complex is considered a growing public health problem due to the negative consequences in the short and long term on people’s well-being (Campo-Arias et al., 2014; Gulliford, 2019).

Consequently, Krieger et al. (2005) carried out the process of construction and validation of the Experiences of Discrimination (EOD) in order to quantify the experiences of discrimination on ethnic-racial grounds in a sample of 228 African Americans and Latinos between 25 and 64 years, residents of the United States. The EOD is made up of nine items that are scored from zero (never) to three (four times or more) and measure the frequency with which an individual has perceived stigma-discrimination in nine contexts: at school, when applying for employment, in work, at home, when renting a house, receiving medical attention, applying for a loan, on the street, and by judicial authorities. The EOD showed a one-dimensional internal structure with acceptable indicators of goodness-of-fit (chi-square = 200.5, degree of freedom = 80, *p* = .05, RMSEA = .05, and CFI = .94) and an acceptable internal consistency (Cronbach’s alpha of .74) (Krieger et al., 2005). In Colombia, Campo-Arias et al. (2014) explored the EOD structure and internal consistency in a group of 361 medical students from Bucaramanga. They reported a one-dimensional internal structure that explained 52.9% of the variance, using exploratory factor analysis (EFA) and high indicators reliability (Cronbach’s alpha of .88 and McDonald’s omega of .89).

Although the EOD was constructed to measure experiences of discrimination by ethnicity or race, the wording of the items allows modification for experiences of discrimination by other characteristics (Krieger et al., 2005). Therefore, Lee et al. (2016) adapted six of the nine items for experiences of discrimination based on sexual orientation during the last year: when obtaining insurance coverage or receiving medical attention, in public places, obtaining a job, work, being admitted to an educational institution and in courts or by the police. However, they did not report any psychometric performance indicators due to the study’s objectives. Subsequently, in Macedonia, Stojanovski et al. (2017) reported that EOD showed high internal consistency (Cronbach’s alpha of .82) in 208 adults of sexual minorities and 188 adults of hegemonic sexual identity.

Therefore, in the present study, a specific adaptation was carried out to know the EOD’s performance by sexual orientation (SOEOD-9) in people with diverse sexual identities, lesbians, gays, bisexuals, and travesties, transgender, transsexual, intersexual, and queer (LGBTTTIQ). LGBTTTIQ people tell a long history of stigma-discrimination based on non-hegemonic sexual identity (Cassey et al., 2019; Wang et al., 2019).

Besides, nomological validity, also known as a hypothesis or theoretical validity, is explored, which seeks to observe the presence or absence of correlation between the construct evaluated by a scale and another independent construct, which theoretically has a statistically significant association (Adcock & Collier, 2001). The present study used the variable “coming out” with the parents to explore the nomological validity. When the individual assumes sexual orientation before the family, it is more frequent for them to behave publicly as a sexually diverse person, and consequently, the risk of discrimination in different socialization contexts increases (Doan Van et al., 2019; Gattamorta & Quidley -Rodríguez, 2018; Wax et al., 2018).

Evidence indicates that experiences of discrimination based on sexual orientation increase the risk of mental health problems and disorders in LGBTTTIQ, including depressive symptoms of anxiety and the spectrum of post-traumatic stress disorder with a significant deterioration in the quality of life (Bialer & McIntosh, 2017; Mason et al., 2018; Petruzzella et al., 2020; Richardson & King, 2017). However, little is known about discrimination experienced in the population with diverse Colombian sexual orientations (Nieves-Lugo et al., 2020). Knowing the implications of discrimination in LGBTTTIQ people requires measurement instruments, such as the SOEOD-9, that can be used in population studies and allow the valid and reliable measurement of experiences of discrimination (González-Rivera & Pabellón-Lebrón, 2018). This knowledge will allow affirmative actions to reduce the impact of discrimination based on sexual orientation and promote the health and well-being of LGBTTTIQ (Lee & Kanji, 2017; Puckett et al., 2016).

This study aimed to know the structure and internal consistency of SOEOD-9 in a group of LGBTTTIQ from Santa Marta, Colombia.

## Method

### Study design and ethical issues

A validation study of a scale was carried out. According to Resolution 8430 of 1993 of the Ministry of Health and Social Protection, the research did not represent any risk for the participants; techniques and methods were applied without intervening or modifying psychological variables anonymously confidentially. Participants must sign an informed consent.

### Participants

Three hundred three individuals with diverse sexual orientations participated, selected by non-probabilistic, snowball-type sampling. The participants’ ages ranged between 18 and 70 years (M = 25.7, SD = 7.3). More information is available in Table 1.

**Table 1.** Characteristic of the sample.

| *Variable* | *Category* | *n (%)* |
| --- | --- | --- |
| **Gender** | Female | 157 ((51.82) |
|  | Male | 140 (46.20) |
|  | Non-binary | 6 (1.98) |
| **Income** | Low | 157 ((51.82) |
|  | Middle | 134 (44.22) |
|  | High | 12 (3.96) |
| **Occupation<sup>1</sup>** | Study | 180 (59.41%) |
|  | Work | 171 (56.44%) |
| **Sexual orientation** | Bisexual | 128 (42.24%) |
|  | Gay | 100 (33%) |
|  | Lesbian | 61 (20.14%) |
|  | Queer | 14 (4.62%) |
| **Parent coming out of the closet** | Yes | 170 (56.11%) |
|  | No | 133 (43.89%) |

<sup>1</sup>Total is higher than 100%.

### Instrument

Participants completed SOEOD-9. The SOEOD-9 comprises nine items that explore discrimination events in nine contexts, from school to interaction with police and authorities.

Each item offers four response options that are scored from zero to three, where zero is “never,” one is “once,” two is “two or three times,” and three is “four times or more.” The version used appears in Table 2.

**Table 2.** SOEOD-9.

| Have you ever experienced discrimination, been prevented from doing something, or been hassled or made to feel inferior in any of the following situations because of your sexual orientation? | Never | Once | 2-3 times | 4 or more times |
| --- | --- | --- | --- | --- |
| 1. At school | 0 | 1 | 2 | 3 |
| 2. Getting a job | 0 | 1 | 2 | 3 |
| 3. At work | 0 | 1 | 2 | 3 |
| 4. Getting housing | 0 | 1 | 2 | 3 |
| 5. Getting medical care | 0 | 1 | 2 | 3 |
| 6. Getting services in store or restaurant | 0 | 1 | 2 | 3 |
| 7. Getting credit, bank loans, or a mortgage | 0 | 1 | 2 | 3 |
| 8. On the street or in a public setting | 0 | 1 | 2 | 3 |
| 9. From the police or in the courts | 0 | 1 | 2 | 3 |

Krieger, N., Smith, K., Naishadham, D., Hartman, C., & Barbeau, E. M. (2005). Experiences of discrimination: validity and reliability of a self-report measure for population health research on racism and health. *Social Science & Medicine*, 61(7), 1576-1596.

### Procedure

The information was collected in person. The printed version consisted of the informed consent, a sociodemographic questionnaire, and the SOEOD-9. The participants filled out the questionnaire anonymously, and each one was assigned a code to preserve confidentiality. The information collected was digitized into a database.

### Analysis of data

The statistical programs IBM-SPSS version 25 and STATA were used. The Bartlett (1950) test of sphericity was performed, in which a high square-chi and probability values (*p*) less than 5% were expected. Likewise, the Kaiser-Meyer-Olkin, KMO (Kaiser, 1974) sample adequacy test was applied, in which a value greater than 0.70 was expected. Adequate indicators in both measures reveal one or more latent factors in a set of items. With the maximum likelihood method and Promax rotation, EFA was used to explore the structure of SOEOD-9, and the communalities and loadings were observed.

In the confirmatory factor analysis (CFA), the goodness-of-fit coefficients were found with the chi-square test, with degrees of freedom (df) and p-value, Root Mean Square Error of Approximation (RMSEA) and confidence intervals of 90% (90%CI), Comparative Fit Index (CFI), Tucker-Lewis index (TLI), and Standardized Mean Square Residual (SRMR). A probability value greater than 5% is expected in the chi-square, with values less than 0.06 for RMSEA and SRMR, and greater than .89 for CFI and TLI. At least three of these indicators were expected to be within desirable parameters (Hu & Bentler, 1999). Other versions would be tested if this were not achieved by observing the modification indices’ results.

The scale’s internal consistency was calculated using the coefficients Cronbach’s alpha of Cronbach and McDonald’s omega of McDonald. Cronbach’s alpha is the internal consistency measure most used in the research; however, McDonald’s omega is a more accurate measure when the tau equivalence principle is not fulfilled, necessary for the precise calculation of internal consistency (Campo-Arias & Oviedo, 2008). It is recommended to report both coefficients to have at least two reliability indicators in the validation studies. Cronbach’s alpha and McDonald’s omega values higher than .70 are expected (Campo-Arias & Oviedo, 2008).

The authors compared the scores on the SOEOD-5 between the participants who reported coming out to their parents and those who did not, using the Student’s t-test (1908) after verifying the homogeneity of the variance with the Levene test (O’Neil & Mathews, 2002). Probability values less than 1% (*p* < .01) were accepted as significant.

## Results

### Dimensionality

The SOEOD-9 showed scores between 0 and 18 (M = 3.64, SD = 3.87). Barlett’s sphericity test indicated a chi-square of 654.9; df = 36, *p* < .001, and the KMO test was .80. These values allowed us to advance in the EFA, in which two latent factors were evidenced: the first with an eigenvalue of 3.4 that explained 37.9% of the variance, which was preliminarily called “proximal discrimination” (items 1, 6, 8, and 9), and the second one called “distal discrimination” (items 2, 3, 4, 5 and 7), with an eigenvalue of 1.2 responsible for 13.7% of the total variance. The subsections showed communalities between .272 and .728 and coefficients between .498 and .875. See details in Table 3.

**Table 3.**
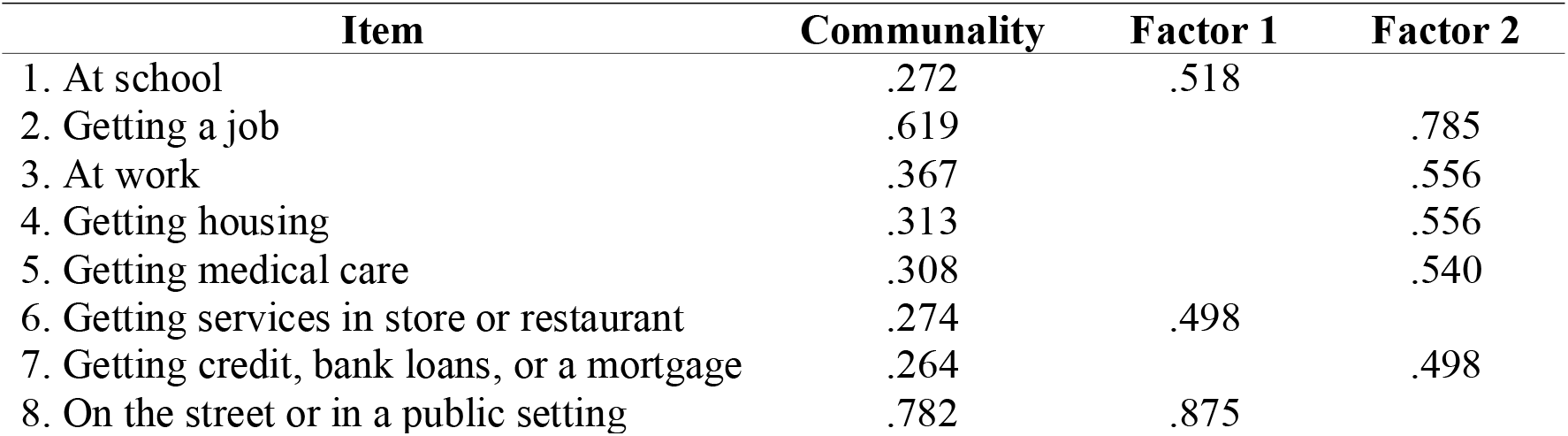
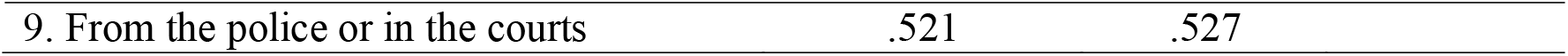
Communalities and loadings of the soeod-9.

| Item | Communality | Factor 1 | Factor 2 |
| --- | --- | --- | --- |
| 1. At school | .272 | .518 |  |
| 2. Getting a job | .619 |  | .785 |
| 3. At work | .367 |  | .556 |
| 4. Getting housing | .313 |  | .556 |
| 5. Getting medical care | .308 |  | .540 |
| 6. Getting services in store or restaurant | .274 | .498 |  |
| 7. Getting credit, bank loans, or a mortgage | .264 |  | .498 |
| 8. On the street or in a public setting | .782 | .875 |  |

|  |  |  |
| --- | --- | --- |
| 9. From the police or in the courts | .521 | .527 |

In the CFA, the goodness-of-fit indicators were poor for the two-dimensional solution (chi-square = 104.54, df = 26, *p* < .001, RMSEA = .10, 90%CI .08 - .12, CFI = .88, TLI = .83, and SRMR = .06). Given these findings, a one-dimensional model was tested with the nine items that make up the scale. Likewise, this solution showed modest goodness-of-fit coefficients (chi-square = 141.18, df = 27, *p* = .001, RMSEA = .12, 90%CI .10 - .14, CFI = .81, TLI = .74, and SRMR = .07).

However, other versions of five items were explored due to the internal consistency coefficients presented below, and the best performance was shown by the version that included items 3, 5, 6, 8, and 9 (SOEOD-5). The SOEOD-5 presented scores between 0 and 12 (M = 2.17; SD = 2.68). The Bartlett test of sphericity showed a chi-square of 288.03; df = 10, *p* < .001, and the KMO test of .795. A single factor with an eigenvalue of 2.5 was retained, which explained 49.4% of the total variance. The goodness-of-fit coefficients were chi-square = 8.11, df = 5, *p* = .15, RMSEA = .05, 90%CI .01 - .10, CFI = .99, TLI = .98, and SRMR = .02. See Table 4 for communalities and coefficients of the SOEOD-5.

**Table 4.** Communalities and loading of the SOEOD-5.

| Item | Communality | Loading |
| --- | --- | --- |
| 1. At work | .499 | .558 |
| 2. Getting medical care | .499 | .556 |
| 3. Getting services in store or restaurant | .525 | .636 |
| 4. On the street or in a public setting | .541 | .656 |
| 5. From the police or in the courts | .505 | .621 |

### Internal consistency

In the one-dimensional model, the SOEOD-9 showed Cronbach’s alpha of 0.75 and McDonald’s omega and .79; and in the two-dimensional model, Cronbach’s alpha was .66 for Factor 1, and Cronbach’s alpha of .70 for Factor 2; and McDonald’s omega of .71 for Factor 1 and McDonald’s omega of .73 for Factor 2. The SOEOD-5 presented Cronbach’s alpha of .71 and McDonald’s omega of .74.

### Nomological validity

The scores on the SOEOD-5 in those who said they had informed their parents of their sexual orientation showed higher scores than those who did not (M = 2.70, SD = 2.85 vs M = 1.48, SD = 2.26, t = 4.15, df = 300.99, *p* < .01). The t value was taken for non-homogeneous variances (F = 11.97, *p* < .001).

## Discussion

In the present study, among LGBTTTIQ people of Santa Marta, Colombia, the SOEOD-9 shows an internal structure of two factors with unacceptable goodness-of-fit coefficients; however, it presents high internal consistency. The SOEOD-5 shows better overall performance with good structure and internal consistency coefficients.

The present investigation observed that the SOEOD-9 presents high internal consistency for one- and two-dimensional structures. However, the goodness-of-fit indicators for these structures are below what is accepted. In contrast, the SOEOD-5 showed a one-dimensional structure with excellent goodness-of-fit indicators and high internal consistency. The SOEOD-5 is the result of eliminating four items with poor performance: item 1 (school), item 2 (applying for a job), item 4 (renting a house), and item 7 (bank loan).

The performance of SOEOD versions is not known with certainty; to date, there is no exploration similar to that of the present study. In Macedonia, Stojanovski et al. (2017) found Cronbach’s alpha of .82, apparently for the SOEOD-9. In ideal conditions, one-dimensional scales are preferred, and the retained factor explains at least 50% of the variance (Campo-Arias et al., 2017; Gorsuch, 1997). Besides, it is expected to observe the goodness-of-fit coefficients within the range preset as acceptable, previously noted (Hu & Bentler, 1999). Finally, it is necessary to have adequate reliability, and for this, it is expected to find internal consistency values between .70 and .95 for Cronbach’s alpha and McDonald’s omega (Campo-Arias & Oviedo, 2008).

Regarding the nomological validity, a higher perception of discrimination was found in the participants who had come out of the closet with their parents. These findings are consistent with the available evidence. Keeping a secret sexual orientation is often used to avoid discrimination among adolescents and young adults (Dueñas et al., 2022; Zheng et al., 2020). Public expression of sexual diversity significantly improves psychological well-being (Perales et al., 2020); however, visibility increases the risk of discrimination, especially in hostile sociocultural contexts toward sexual diversity (Suppes et al., 2021; Wei et al., 2017).

The SOEOD-5 is a short, valid, and reliable measure that quantifies discrimination experiences based on sexual orientation. The best overall performance was observed for the SOEOD-5 in the present investigation. This observation is promising, given that it allows us to have an instrument for measuring the experience of discrimination based on sexual orientation in a diverse Colombian sexual population (González-Rivera & Pabellón-Lebrón, 2018). These findings constitute a significant empirical contribution to the medical and social sciences and research that explores the experiences of discrimination based on sexual orientation and mental health in the LGBTTTIQ population.

The present study makes a notable contribution to scientific knowledge; however, it is necessary to recognize some research limitations. Homosexuality and bisexuality are still taboo subjects in Colombian society (Zambrano et al., 2019), making it difficult to approach the study population. At the time of application, only people who publicly recognized, to a greater or lesser extent, diverse sexual orientation was included in the sample. The psychometric performance of the different versions of SOEOD can show different indicators if the individuals who remain “in the closet” are evaluated. It is possible that in this group of people, a very distant performance is observed, and, consequently, the results should be interpreted with caution and can only be reproduced in people who are publicly self-recognized as LGBTTTIQ (Baams et al., 2013).

Exploring the specific psychometric performance of the SOEOD-5 in each sexual orientation was not recommended due to the small groups formed with the LGBTTTIQ group’s segmentation. Caution should be exercised because sociocultural factors moderate the manifestation of discriminatory behaviors, which vary according to sexual orientation, identity, and gender expression (Balsam et al., 2013; Cerezo, 2020).

It is concluded that the SOEOD-9 is an instrument with adequate indicators of internal consistency and two factors with imperfect goodness-of-fit (proximal discrimination and distal discrimination). The SOEOD-5 shows better overall performance, both in dimensionality and internal consistency. It is necessary to carry out other studies to know the psychometric performance of different versions of SOEOD.

## Data Availability

The data supporting this study's findings are available from the corresponding author upon reasonable request.

## References

Adcock, R., & Collier, D. (2001). Measurement validity: A shared standard for qualitative and quantitative research. American Political Science Review, 95(3), 529–546. https://www.jstor.org/stable/3118231

Allport, G. (1954). The nature of prejudice. Addison-Wesley Publishing Company.

Awad, G., & Reckley, K. (2017). The International Encyclopedia of Intercultural Communication (K. Young). John Wiley & Sons, Inc.

Baams, L., Beek, T., Hille, H., Zevenbergen, F., & Bos, H. (2013). Gender nonconformity, perceived stigmatization, and psychological well-being in Dutch sexual Gay-specific and general stressors predict gay men’s psychological functioning over time minority youth and young adults: A mediation analysis. Archives of Sexual Behavior, 42(5), 765–773. https://doi.org/doi:10.1007/s10508-012-0055-z

Balsam, K., Beadnell, B., & Molina, Y. (2013). The Daily Heterosexist Experiences Questionnaire. Measurement and Evaluation in Counseling and Development, 46(1), 3–25. https://doi.org/10.1177/0748175612449743

Bartlett, M. (1950). Tests of significance in factor analysis. British Journal of Statistical Psychology, 3(2), 77–85. https://doi.org/10.1111/j.2044-8317.1950.tb00285.x

Bialer, P., & McIntosh, C. (2017). Discrimination and LGBT mental health. Journal of Gay & Lesbian Mental Health, 21(4), 275–276. https://doi.org/10.1080/19359705.2017.1356138

Campo Arias, A., & Herazo, E. (2013). Estigma, prejuicio y discriminación en salud mental [Stigma, prejudice and discrimination in mental health]. Revista Ciencias Biomédicas, 4(1), 9–10.

Campo-Arias, A., & Herazo, E. (2014). Estigma y salud mental en personas víctimas del conflicto armado interno colombiano en situación de desplazamiento forzado [Stigma and mental health in victims of Colombia’s internal armed conflict in situation of forced displacement]. Revista Colombiana de Psiquiatría, 43(4), 212–217. https://doi.org/10.1016/j.rcp.2014.09.004

Campo-Arias, A., & Herazo, E. (2015). El complejo estigma-discriminación asociado a trastorno mental como factor de riesgo de suicidio [The stigma-discrimination complex associated with mental disorder as a risk factor for suicide]. Revista Colombiana de Psiquiatría, 44(4), 243–250. https://doi.org/10.1016/j.rcp.2015.04.003

Campo-Arias, A., & Oviedo, H. (2008). Propiedades psicométricas de una escala: la consistencia interna [Psychometric properties of a scale: internal consistency]. Revista de Salud Pública, 10(5), 831–839. https://doi.org/10.1590/S0124-00642008000500015

Campo-Arias, A., Herazo, E., & Oviedo, H. C. (2012). Análisis de factores: fundamentos para la evaluación de instrumentos de medición en salud mental [Factor analysis: principles to evaluate measurement tools for mental health]. Revista Colombiana de Psiquiatría, 41(4), 659–671. https://doi.org/10.1016/S0034-7450(14)60036-6

Campo-Arias, A., Oviedo, H. C., & Herazo, E. (2014). Escala de experiencias de discriminación: Consistencia y estructura interna en estudiantes de medicina [Experiences of Discrimination Scale: Internal consistency and structure in medical students]. CES Psicología, 7(2), 15–26.

Carter, R., Johnson, V., Kirkinis, K., Roberson, K., Muchow, C., & Galgay, C. (2019). A meta-analytic review of racial discrimination: relationships to health and culture. Race and Social Problems, 11(1), 15–32. https://doi.org/10.1007/s12552-018-9256-y

Carter, R., Lau, M., Johnson, V., & Kirkinis, K. (2017). Racial discrimination and health outcomes among racial/ethnic minorities: A metalJanalytic review. Journal of Multicultural Counseling and Development, 45(4), 232–259. https://doi.org/10.1002/jmcd.12076

Casey, L., Reisner, S., Findling, M., Blendon, R., Benson, J., Sayde, J., & Miller, C. (2019). Discrimination in the United States: Experiences of lesbian, gay, bisexual, transgender, and queer Americans. Health Services Research, 54(2), 1454–1466. https://doi.org/10.1111/1475-6773.13229

Cerezo, A. (2020). Expanding the reach of Latinx psychology: Honoring the lived experiences of sexual and gender diverse Latinxs. Journal of Latinx Psychology, 8(1), 1–6. http://dx.doi.org/10.1037/lat0000144

De Freitas, D., Fernandes-Jesus, M., Ferreira, P., Coimbra, S., Teixeira, P., de Moura, A., Gato, J., Marques, S., & Fontaine, A. (2018). Psychological correlates of perceived ethnic discrimination in Europe: A meta-analysis. Psychology of Violence, 8(6), 712–725. https://doi.org/10.1037/vio0000215

Doan Van, E., Mereish, E., Woulfe, J., & Katz-Wise, S. (2019). Perceived discrimination, coping mechanisms, and effects on health in bisexual and other non-monosexual adults. Archives of Sexual Behavior, 48(1), 159–174. https://doi.org/10.1007/s10508-018-1254-z

Dueñas, J., Morales-Vives, F., & Galea, N. (2022). Psychological issues among Spanish adolescents and young people when coming out of the closet to their families. Psychological Reports, 332941211069518. https://doi.org/10.1177/00332941211069518

Gattamorta, K., & Quidley-Rodriguez, N. (2018). Coming out experiences of Hispanic sexual minority young adults in South Florida. Journal of Homosexuality, 65(6), 741–765. https://doi.org/10.1080/00918369.2017.1364111

Ghumman, S., & Ryan, A. (2018). Oxford library of psychology. The Oxford handbook of workplace discrimination (A. Colella, & E. King Eds.). Oxford University Press.

Goffman, E. (1963). Stigma: Notes on the management of spoiled identity. Simon and Schuster.

González-Rivera, J., & Pabellón-Lebrón, S. (2018). Desarrollo y validación de un instrumento para medir discriminación percibida en la comunidad LGBT [Development and validation of an instrument for the measurement of perceived discrimination in the LGBT community]. Revista Evaluar, 18(2), 59–74. https://doi.org/10.35670/1667-4545.v18.n2.20809

Gorsuch, R. L. (1997). Exploratory factor analysis: Its role in item analysis. Journal of Personality Assessment, 68(3), 532–560. https://doi.org/10.1207/s15327752jpa6803_5

Gulliford, M. (2019). Discrimination and public health. The Lancet Public Health, 4(4), e173–e174. https://doi.org/10.1016/S2468-2667(19)30044-1

Heilman, M., & Caleo, S. (2018). Combatting gender discrimination: A lack of fit framework. Group Processes & Intergroup Relations, 21(5), 725–744. https://doi.org/10.1177/1368430218761587

Hendry, J. (2016). An introduction to social anthropology: sharing our worlds. Palgrave.

Hernández, A., & López, M. (2011). Bases teóricas del estigma, aproximación en el cuidado de personas con herpes genital [Theoretical approaches of stigma, analysis in persons with genital herpes]. Index de Enfermería, 20(3), 174–178. http://dx.doi.org/10.4321/S1132-12962011000200008

Hu, L. T., & Bentler, P. M. (1999). Cutoff criteria for fit indexes in covariance structure analysis: Conventional criteria versus new alternatives. Structural Equation Modelling, 6(1), 1–55. https://doi.org/10.1080/10705519909540118

Jones, K., Sabat, I., King, E., Ahmad, A., McCausland, T., & Chen, T. (2017). Isms and schisms: A metalJanalysis of the prejudicelJdiscrimination relationship across racism, sexism, and ageism. Journal of Organizational Behavior, 38(7), 1076–1110. https://doi.org/10.1002/job.2187

Kaiser, H. (1974). An index of factorial simplicity. Psychometrika, 39(1), 31–36. https://doi.org/10.1007/BF02291575

Krieger, N., Smith, K., Naishadham, D., Hartman, C., & Barbeau, E. M. (2005). Experiences of discrimination: validity and reliability of a self-report measure for population health research on racism and health. Social Science & Medicine, 61(7), 1576–1596. https://doi.org/10.1016/j.socscimed.2005.03.006

Lee, A., & Kanji, Z. (2017). Queering the health care system: Experiences of the lesbian, gay, bisexual, transgender community. Canadian Journal of Dental Hygiene, 51(2), 80–89.

Lee, J. H., Gamarel, K. E., Bryant, K. J., Zaller, N. D., & Operario, D. (2016). Discrimination, mental health, and substance use disorders among sexual minority populations. LGBT Health, 3(4), 258–265. https://doi.org/10.1089/lgbt.2015.0135

Lefevor, G. T., Park, S. Y., Acevedo, M. J., & Jones, P. J. (2020). Sexual orientation complexity and psychosocial/health outcomes. Journal of Homosexuality (ahead of print). https://doi.org/10.1080/00918369.2020.1815432

Lippmann, W. (1991). Public opinion. Transaction Publishers.

López, E., Berrios, P., & Augusto, J. (2008). Introducción a la psicologia social [Introduction to social psychology]. Colección Universitas.

Mason, S., Doane, M., & Elliott, M. (2018). Gay and lesbian experiences of discrimination, health, and well-being: Surrounding the presidential election. Social Psychological and Personality Science, 9(2), 131–142. https://doi.org/10.1177/1948550617732391

Ministerio de Salud [Ministry of Health] (1993). Por la cual se establecen las normas científicas, técnicas y administrativas para la investigación en salud. [By which the scientific, technical, and administrative standards for health research are established].

Nieves-Lugo, K., Barnett, A., Pinho, V., Sáenz, M., & Zea, M. C. (2020). Perspectives on sexual orientation and diversity. LGBTQ mental health: International perspectives and experiences (N. Nakamura, & C. Logie). American Psychological Association.

O’Neil, M. E., & Mathews, K. L. (2002). Levene tests of homogeneity of variance for general block and treatment designs. Biometrics, 58(1), 216–224.

Perales, F., Simpson Reeves, L., Plage, S., & Baxter, J. (2020). The family lives of Australian lesbian, gay and bisexual people: A review of the literature and a research agenda. Sexuality Research and Social Policy, 17(1), 43–60. https://doi.org/10.1007/s13178-018-0367-4

Petruzzella, A., Feinstein, B. A., Davila, J., & Lavner, J. A. (2020). Gay-specific and general stressors predict gay men’s psychological functioning over time. Archives of Sexual Behavior, 1–13. https://doi.org/10.1007/s10508-020-01672-4

Puckett, J., Newcomb, M., Ryan, D., Swann, G., Garofalo, R., & Mustanski, B. (2016). Internalized homophobia and perceived stigma: a validation study of stigma measures in a sample of young men who have sex with men. Sexuality Research and Social Policy, 14(1), 1–16. https://doi.org/10.1007/s13178-016-0258-5

Ramírez, E., Estrada, C., & Yzerbyt, V. (2016). Estudio correlacional de prejuicio y discriminacion implicita y explicita en una muestra magallanica [Correlational study on implicit and explicit prejudice and discrimination in a Magellan sample]. Atenea, (513), 251–262. http://dx.doi.org/10.4067/S0718-04622016000100016

Richardson, V., & King, S. (2017). Mental health for older LGBT adults. Annual Review of Gerontology and Geriatrics, 37(1), 59–75. https://doi.org/10.1891/0198-8794.37.59

Semlyen, J., King, M., Varney, J., & Hagger-Johnson, G. (2016). Sexual orientation and symptoms of common mental disorder or low well-being: a combined meta-analysis of 12 UK population health surveys. BMC Psychiatry, 16(1), 67. https://doi.org/10.1186/s12888-016-0767-z

Stangor, C. (2016). Handbook of prejudice, stereotyping, and discrimination (T. Nelson). Psychology Press.

Stojanovski, K., Zhou, S., King, E., Gjorgjiovska, J., & Mihajlov, A. (2017). An application of the minority stress model in a non-western context: Discrimination and mental health among sexual and gender minorities in Macedonia. Sexuality Research and Social Policy, 15(3), 367–376. https://doi.org/10.1007/s13178-017-0299-4

Student (1908). The probable error of a mean. Biometrika, 6(1), 1–25. https://doi.org/10.2307/2331554

Sugarman, D. B., Nation, M., Yuan, N. P., Kuperminc, G. P., Hassoun Ayoub, L., & Hamby, S. (2018). Hate and violence: Addressing discrimination based on race, ethnicity, religion, sexual orientation, and gender identity. Psychology of Violence, 8(6), 649–656. http://dx.doi.org/10.1037/vio0000222

Suppes, A., van der Toorn, J., & Begeny, C. T. (2021). Unhealthy closets, discriminatory dwellings: The mental health benefits and costs of being open about one’s sexual minority status. Social Science & Medicine (1982), 285(114286), 114286. https://doi.org/10.1016/j.socscimed.2021.114286

Wang, Y., Hu, Z., Peng, K., Xin, Y., Yang, Y., Drescher, J., & Chen, R. (2019). Discrimination against LGBT populations in China. The Lancet Public Health, 4(9), e440–e441. https://doi.org/10.1016/S2468-2667(19)30153-7

Wax, A., Coletti, K. K., & Ogaz, J. W. (2018). The benefit of full disclosure: A meta-analysis of the implications of coming out at work. Organizational Psychology Review, 8(1), 3–30. https://doi.org/10.1177/2041386617734582

Wei, C., & Liu, W. (2019). Coming out in Mainland China: A national survey of LGBTQ students. Journal of LGBT Youth, 16(2), 192–219. https://doi.org/10.1080/19361653.2019.1565795

Zambrano, C., Hernández, P., & Guerrero, P. (2019). Proceso de reconocimiento de la orientación sexual homosexual en estudiantes de una universidad pública [Recognition process related to homosexual orientation in students of a public university]. Psicogente, 22(41), 243–271. http://dx.doi.org/10.17081/psico.22.41.3310

Zschirnt, E., & Ruedin, D. (2016). Ethnic discrimination in hiring decisions: a meta-analysis of correspondence tests 1990–2015. Journal of Ethnic and Migration Studies, 42(7), 1115–1134. https://doi.org/10.1080/1369183X.2015.1133279

